# Understanding the evidence for climate concerns, negative emotions and climate-related mental ill-health in young people: a scoping review

**DOI:** 10.1101/2021.09.27.21264151

**Authors:** Reem Ramadan, Alicia Randell, Suzie Lavoie, Caroline X Gao, Paula Cruz Manrique, Rebekah Anderson, Isabel Zbukvic

## Abstract

**Background:** Human-induced climate change represents a serious threat to human health, including mental health, due to both the traumatising effects of extreme climate events and the psychological effect of worry about climate change. The present scoping review aims to systematically search and synthesise original research related to mental health and climate related concerns, negative emotions and mental ill-health in young people. Findings will help to understand the current landscape, gaps in the literature, and to provide recommendations for future youth mental health research and practice.

**Methods:** A systematic search and narrative synthesis of the literature published prior to October 2020 examining negative emotions associated with climate change in young people was undertaken. Studies were included if they examined mental ill-health (e.g. symptoms of depression or anxiety) or negative emotions (e.g. distress, worry, concern) associated with the threat of climate change.

**Findings:** Of the 3329 peer-reviewed articles screened, 12 met the inclusion criteria. Together, included studies show that young people are worried about climate change. Studies also explored the coping strategies young people use to manage their distress caused by climate change.

**Interpretations:** The limited literature in this area indicates a key gap in youth mental health research. Available evidence suggests that young people are concerned about climate change, which may increase risk of mental ill-health. Thus, clinicians should assess for and address climate anxiety in young people. Mental health leaders are urged to advocate for urgent climate action to mitigate the effects of climate anxiety in young people.

**Funding:** this project was not funded.

## INTRODUCTION

Climate change has been described as the biggest threat to global health of the 21^st^ century.^1^ In addition to having a deleterious impact on a range of physical health outcomes, climate change also poses a major threat to mental health,^2^ referring not only to mental illnesses or disorders, but to general wellbeing, cognition, and emotional states.^3^ Several reviews have been published in recent years that aimed to synthesise the existing literature on climate change and mental health.^4-8^ These reviews based on a large body of research show that climate change can negatively impact mental health via exposure to traumatic events such as drought or hurricane. However, relatively less attention has been given to understanding the mental health impacts of worry or concern about climate change. Table 1 provides a summary of terms used to describe negative emotions related to climate change (e.g., worry, distress, fear or despair) from existing literature.

**Table 1.** Concepts of negative emotions related to environment and climate change.

| Term | Definition |
| --- | --- |
| Climate (change) anxiety<sup>10</sup> | The experience of intense anxiety associated with perceptions about climate change, even among people who have not personally experienced substantial adverse impacts of climate change. |
| Climate change worry<sup>14</sup> | Trouble, disturbing thoughts that people experience about climate change. |
| Eco-anxiety<sup>15</sup> | As people become overwhelmed by scale and complexity of climate change and other issues with ecosystem, they become anxious about what to do in the face of accumulating evidence of non-sustainable pathways. This in turn leads to fear, concern, and despair for the individual's own future, that of their children and later generations. |
| Eco-anxiety<sup>16</sup> | A chronic fear of environmental doom. |
| Ecological grief<sup>17</sup> | The grief felt in relation to experienced or anticipated ecological losses, such as the loss of species, ecosystems and meaningful landscapes due to acute or chronic environmental change. |
| Ecological stress<sup>18</sup> | Perceptions of stress related to a changing and uncertain environment. |
| Ecoparalysis<sup>15</sup> | Feelings of powerlessness and inability to act and respond to the climatic and ecological challenges. |
| Environmental distress<sup>19</sup> | Concept of multi-stage process including perceiving environmental changes, identifying threats, developing emotional reaction and endorsing actions or adaptations. |
| Environmental grief<sup>20</sup> | The grief reaction stemming from the environmental loss of ecosystems caused by natural or human-made events. |
| Solastalgia<sup>21</sup> | Distress that is produced by environmental change impacting on people while they are directly connected to their home environment. |

Although worrying about climate change is not pathological by itself, negative climate-related emotions can become clinically significant when they grow to being difficult to control.^9-11^ Experiences of distress and anxiety related to climate change may also exacerbate other mental health problems or compound daily stressors to negatively affect overall mental health, wellbeing, quality of life and functioning.^12,13^ Understanding how climate anxiety and related experiences can impact mental health is critical for understanding the full impacts of climate change on human health.

Several reviews of the literature have highlighted young people as a key group likely to be disproportionately affected by the mental health effects of climate change.^4,22^ Childhood, adolescence and young adulthood are seen as critical periods of development in terms of mental ill-health, with 75% of mental illnesses emerging before the age of 25 years.^23,24^ Experiencing mental ill-health during youth can have significant and lasting impacts, affecting mental health, wellbeing, and functional outcomes over the lifetime. Globally, youth-led climate action as well as the evidence from grey literature suggests that young people are concerned about climate change.^25-27^ Therefore, understanding the short- and long-term mental health effects of negative emotions related to climate change in young people is key to supporting positive outcomes for individuals, communities, and society. The Lancet recently published a call to action related to climate anxiety in young people, which emphasised the urgent need for further research in this area.^28^ However, there are currently no known peer-reviewed literature reviews focused on the evidence related to mental health and negative emotions about climate change in young people.

The present scoping review aims to systematically search and synthesise original research related to mental health and climate-related concerns, negative emotions and mental ill-health in young people. Findings will help to understand the current landscape, gaps in the literature, and to provide recommendations for future youth mental health research and practice.

## METHODS

This review was conducted according to the Preferred Reporting Items for Systematic Reviews and Meta-Analyses extension for Scoping Reviews (PRISMA-ScR) guidelines.^29,30^ The protocol (unpublished) for the full search strategy can be seen in Supplementary Materials. Computerised literature searches of Embase, PsycINFO and MEDLINE were conducted in October 2020. The searches did not apply any limitation on the year of publication or study design. The reference lists of included studies and reviews were also manually searched for any additional articles for potential inclusion. Articles selected were screened initially by title and abstract by AR, RA and IZ. Articles flagged as potentially fulfilling the inclusion criteria were retrieved, and full text screening was completed by AR, RA, IZ, RR and SL. Full text screening assessed articles for inclusion against the inclusion criteria.

Studies were included in this review if they met the following criteria: 1) the study reported primary research; 2) the paper reported negative emotions (e.g., worry, concern) and/or mental ill-health in relation to the threat of climate change; 3) the sample or a subset of the sample was age 26 years or younger, or described as young people, teens, students, adolescents, children, or similar; 4) the paper was written in English, French or German; 5) the paper was published in a peer-reviewed journal. Articles were excluded if they reported negative emotions and/or mental ill-health following a natural disaster or in response to a specific climatic event or causative factor, as this review aimed to examine the psychological presentations resulting from concerns related to changing climate or environment. Studies that measured a correlation between mental health and a specific weather or environmental factors (e.g., air temperature, level of air pollution) were therefore excluded. Any studies that were in doubt for inclusion were discussed by the authors until consensus was reached.

Ten percent of the full text results were randomly selected and double-screened by PCM. PCM was blind to the initial inclusion/exclusion status of studies. Any discrepancies were resolved through discussion in team meetings, and by making the inclusion and exclusion criteria clearer. In this initial 10% of full text articles that were double screened, the discrepancies revealed indicated that the inclusion criteria were not strict enough resulting in ineligible studies being included by PCM, as opposed to eligible studies being excluded. Consequently, the authors concluded that it was unlikely any eligible studies had been screened out.

The following data were extracted from included studies into a standardised data collection spreadsheet: number of young people included in the study, their mean age or age range, the country where the study was conducted as well as type of sample selected, data type and collection methods, climate-specific mental health outcome and findings, factors associated with climate anxiety and worry, protective and coping factors, and authors’ key conclusion. Study findings were summarised by the types of outcomes they examined, the country where research was conducted, and whether there was an older comparison group.

Although not the focus of this review, literature screening also included consideration of articles that examined the effect of natural disasters on youth mental health. Any article screened that discussed the effect of a natural disaster on the mental health of young people was compiled in a separate list to examine trends in number of publications over time.

## RESULTS

Figure 1 shows that 3329 articles were considered for inclusion in this scoping review, 174 full-text articles were assessed for eligibility and 12 records fulfilled the inclusion criteria. Articles were excluded mainly due to the study participants not being young people, the articles being reviews or opinion pieces rather than original research, outcomes not being related to mental health, or mental health outcome/s being linked to climate change via an extreme weather event. Included studies were published between 1995 and 2019 and were undertaken across five countries, all developed countries. Table 2 summarises key details of included studies.

**Table 2.** Details of Studies that Investigated the Effect of Climate Anxiety on Young People’s Mental Health

| First author (year) | No. of young people | Mean age in years or age range | Country (sample) | Data type (collection methods) | Climate-specific mental health outcome (measure) | Climate-specific mental health findings | Factors associated with climate anxiety or worry | Protective and coping factors | Key conclusion by authors |
| --- | --- | --- | --- | --- | --- | --- | --- | --- | --- |
| Henker, Whalen & O’Neil (1995)<sup>31</sup> | 194 | Grade 4 – 8 | USA (public USA schools) | Qualitative and quantitative (structured interview) | Worry profiles: health and environmental risk perceptions in terms of seriousness and perceived control, prevalence, and personal vulnerability. | 28·4% worried about environmental degradation (e.g., “when I grow up there might be too much pollution to live on this planet”). | Hypothesised link between increased rates of depression and worry about societal issues in females, which can trigger feelings of hopelessness and helplessness. | Gender; male students were less likely to worry more about global and societal issues. | Concerns about environmental degradation are relatively common, expressed by approximately one-quarter of students. |
| Ojala (2005)<sup>32</sup> | 253 | 17·5 | Sweden (high school students) | Quantitative and qualitative (self-report survey) | Worry about environmental risks, 11-item questionnaire. | Overall 26·4% of the participants scored high on environmental worry. Two clusters with adolescents who were high on worry could be identified. One scored high on well-being (11·6% of total sample) and one scored low on the same scale (14·8% of total sample). | Altruistic and biospheric values, as well as high levels of trust in science and environmental organisations are correlated with environmental worry. | Protective factors identified include: meaningfulness, hope, anger, and trust in environmental organisations. | Environmental worry may co-occur with both positive and negative well-being. Young people with higher wellbeing were found to have higher meaningfulness, hope, anger, and trust in environmental organizations. Level of trust held for societal institutions, science, and their own environmental self-efficacy was found similar between those had higher and lower wellbeing. |
| Wolf & Salo (2008)<sup>33</sup> | 1 | 17 | Australia (Major Depressive Disorder (MDD) diagnosis) | Qualitative (case study) | DSM Diagnosis and Level of distress. | Diagnosis of MDD with psychotic features - ‘Climate change delusion’. | High level of concern and distress about climate change. | Not measured | Ideas and beliefs about climate change can manifest as psychotic phenomena, as has been observed with other contemporary phenomena, particularly as climate change concerns become increasingly common. |
| Agho et al. (2010)<sup>34</sup> | 2004 total – however number not reported for the 16-24 group | 16 and over with separate findings for the 16-24 group | Australia (general population, 16 and over) | Quantitative (survey) | Concerns about worsening effects of global warming in Australia and direct impact on self and family (2-items). Psychological distress measured using the K10, with 'high' psychological distress considered as being > 22. | Half of respondents aged 16-24 (50.6%), were concerned that they or their family would be directly affected if global warming were to worsen in Australia. 58.2% thought global warming was likely to worsen and 74.1% said they had changed the way they lived their life because of the possibility of worsening global warming. | Individuals (across entire sample) with high levels of psychological distress were more than twice as likely to think that global warming would worsen compared to individuals with low psychological distress. | Age/generation; older Australians perceived lower risk of increasing global warming. | The majority of young people think that global warming was likely to worsen, expressed concern that they or family members would be adversely affected by this and had changed their way of living due to this possibility. |
| Searle & Gow (2010)<sup>35</sup> | Not reported. Of 275 participants, 173 were first year psychology students | Not reported. 57% of sample aged 18-25 | Australia (university students and members of the general public) | Quantitative (survey) | Climate change distress (12-item)<br>Future change anxiety (29-item)<br>Climate change hopelessness (1-item).<br>Depression, anxiety & stress (DASS-21). | Climate change distress was positively associated with depression, anxiety and stress. | Younger participants (18-25 years) were significantly more distressed about climate change than older (36+ years) participants. Pro-environmental beliefs and high levels of future anxiety and intolerance of uncertainty were also associated with climate change distress measures. | Gender; males were significantly less distressed than females in all climate change measures. | The public is becoming increasingly concerned about climate change and this is associated with negative emotional reactions in certain individuals, particularly younger people. |
| Ojala et al. (2012a)<sup>36</sup> | 348 | Late childhood mean age 11·7<br>Adolescent mean age 16·4<br>Young adulthood mean age 22·6 | Sweden (young people) | Quantitative and qualitative (survey) | Hope concerning climate change (1-item) and worry about climate change (1-item). Coping with worry (3-items). Reasons for hope (1-item). | 29% of children, 62% of adolescents and 62% of young adults indicated feeling worried about climate change fairly much, a lot, or very much. | Not measured | Age: hope about climate change and increased hyperactivation (including rumination and a focus on negative emotions) decreased in younger children. | Young people use a plethora of different strategies to cope with worries about climate change (emotion-focused coping – distancing and individual problem-focused coping strategies) and promote hope (collective problem-focused coping strategies and meaning-focused coping).<br>De-emphasizing the seriousness of the problem was the most common strategy used by people who were not worried about climate change. |
| Ojala et al. (2012b)<sup>37</sup> | 293 | 12 | Sweden (12-year-olds students) | Quantitative (survey) | Optimism concerning climate change (3-item) and worry about climate change (5-item). General negative affect measured by Child Depression Scale. | Association between climate change and negative affect was not directly measured. | Highly problem-focused children worry more about climate change. Problem-focused coping was positively related to general negative affect. | Children who engaged in meaning-focused coping, purpose, and optimism were less likely to experience negative affect and more likely to experience life satisfaction and general positive affect. | The importance of meaning-focused coping and positive emotions for constructive coping with climate change among children is highlighted.<br>Problem-focused coping strategies may not mitigate excessive negative affect from climate change worries, but do positively relate to environmental engagement.<br>Optimism and hope not only provide comfort, but also seem to be a motivational force to environmental engagement. |
| Strife et al. (2012)<sup>38</sup> | 50 | 10 - 12 | USA (children from various ethnic and socioeconomic backgrounds) | Qualitative (In-depth interviews) | Environmental concern; defined as an affective environmental attitude that refers to the emotions people feel about environmental problems, ranging from fear and worry about environmental problems to a general desire to care for the environment | Eighty-two percent of children expressed environmental concern through the emotions of sadness, fear, and anger and frustration in reaction to environmental problems. | Helplessness and feeling that very little is being done by adults and the general public to help solve environmental problems. Pessimistic visions of the future were common, but for some this was dependent on if humans did not change their behaviour. | An awareness of the possible solutions to environmental problems was associated with hope for a better world and hope for positive change. | Many children express their environmental concern through the experience of negative emotions and pessimism about the future. Age-appropriate messaging and education that offers solutions to environmental problems can help mitigate these negative reactions. |
| Ojala et al. (2013)<sup>39</sup> | 321 | 17 | Sweden (senior high school students) | Quantitative (survey) | Optimism concerning climate change (3-items) and worry about climate change (5-items).<br>Life satisfaction (7-items), and anxiety and depressive feelings during last week (general negative affect; 7-items). | Not measured | Worry about climate change mediated the association between problem-focused coping and general negative affect. The more problem-focused coping the adolescents used, the more likely it was that they experienced negative affect in everyday life. | Meaning-focused coping was positively related to both well-being and optimism concerning climate change. | Replicates finding of a previous study on younger children showing that problem-focused and meaning-focused coping were positively related to felt efficacy and environmental behaviour, while de-emphasizing the threat was negatively related to these measures. |
| El Zoghbi & El Ansari (2014)<sup>40</sup> | 66 | 18 - 30 | The Netherlands (University students) | Qualitative (focus groups and interviews) | Concern about ethical implications of climate change. | Not measured | Participants reported feeling guilty and responsible regarding global inequalities leading to disproportional effect of climate change on developing countries. | Not measured | University students' moral concerns and contributions with regards to climate change can have various implications for their short and long-term well-being. Holding an ecological worldview and desire to play a positive role in society, as well as feeling guilty and responsible for perceived inequalities, increased motivation to contribute to change and environmental sustainability. |
| MacDonald et al. (2015)<sup>41</sup> | 17 | 15 – 25 | Canada (Nunatsiavut Inuit youth) | Qualitative (interview) | Feelings about changes in the environment, coping and perceived effect of climate change on mental health. | Changes in climate and environment limited access to protective factors and fostered negative emotions, including worry, boredom, loneliness, isolation, restlessness, fear, sadness, anger, frustration, stress, and anxiety. | Not measured | Six themes identified as protective: being on the land; connecting to Inuit culture; strong communities; relationships with family and friends; staying busy; and adaptability. | The findings are consistent with research in other geographical contexts that points to the significant impacts of climate and environmental change on the mental health and well-being of individual young people and their communities. |
| Li & Monroe (2017)<sup>42</sup> | 728 | Grade 9 - 12 | USA (high school students from south-eastern states) | Quantitative (survey) | Concern about climate change (1-item). | Not measured | Gender (female), race (non-white), and knowing people involved in environmental management (of forests) lead to greater concern about climate change. | Positive association between concern about climate change and hope about ability to influence climate change solutions. Also, being female and older, having opportunities to learn about practice strategies, and knowing people who manage their forests lead to greater hopefulness. | Young people who are strongly concerned about climate change may also experience a higher degree of hope; perhaps greater concern helps trigger greater attention to gaining information and examples and finding strategies to be effective. |

**Figure 1.**
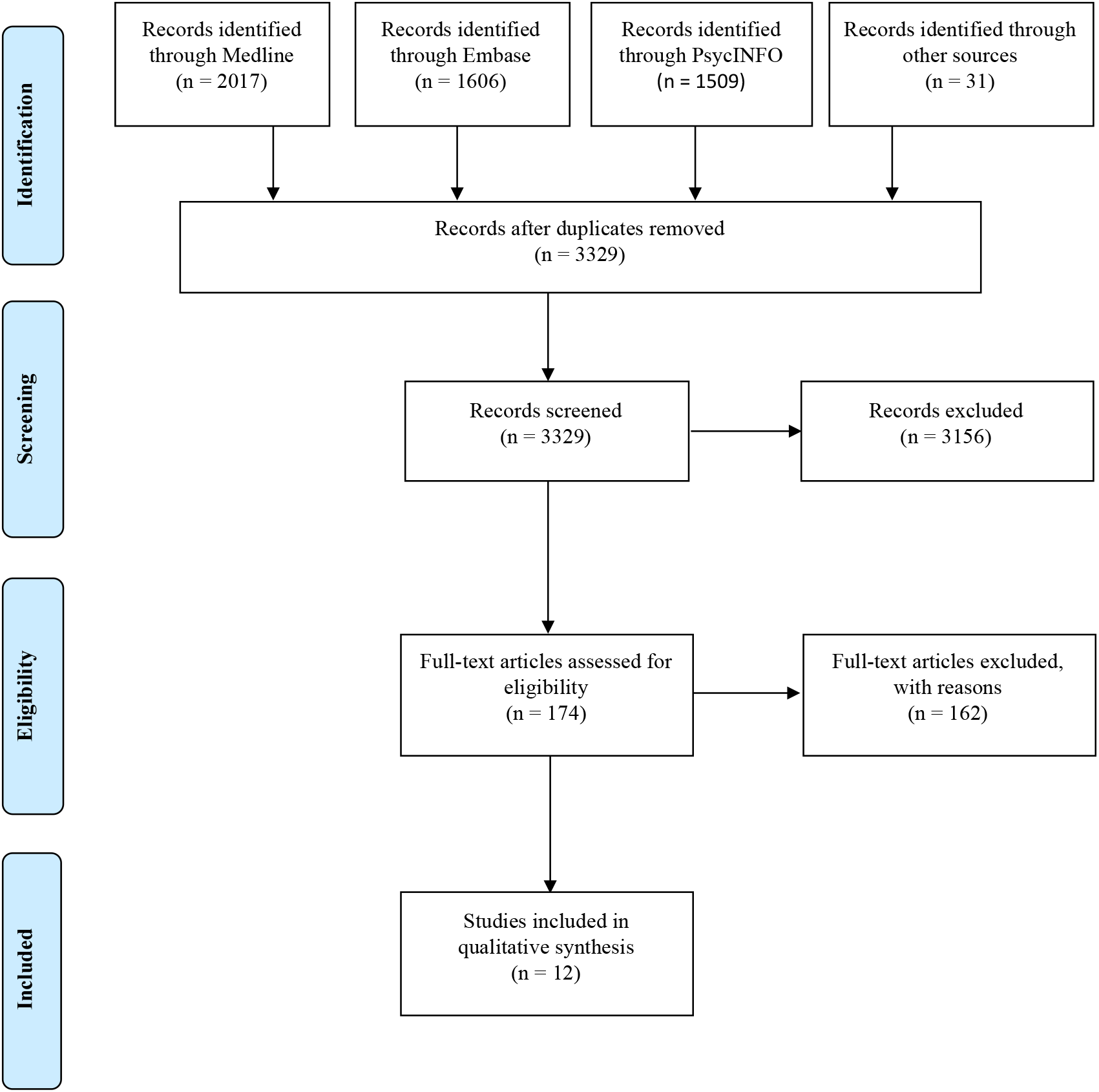
PRISMA-ScR Flow diagram ^29^

### Main analysis

Of the 12 studies included, four studies aimed to provide general understanding of climate concerns or negative emotions in the population,^31,34,35,38^ seven focused on understanding the interplay between climate-related concerns and negative emotions, with psychological distress, wellbeing, hope, as well as coping mechanisms, ^32,36,37,39-42^ and one clinical case report focused on climate change-related psychotic features.^33^ Two studies, both conducted in Australia, reported age-group comparisons and suggested a higher level of climate distress among young people compared with older populations.^34,35^ Studies predominantly used interviews, focus groups, or surveys to collect data from young people, and almost half the studies included in this review were conducted by one research group.

Studies examined a range of negative emotions, mental health outcomes, and coping strategies related to the threat of climate change (Table 3). Most studies focused on worry or concern.^31,32,34,36,37,40,42^ Two studies included a range of different negative emotions such as anger, stress, and boredom,^35,41^ while one investigated self-described feelings.^38^ Notably, most included studies examined factors associated with supporting wellbeing or coping in the face of climate change. Of these, most focused on efficacy (i.e., the belief that individuals and communities can take action) and hope.^36,37,42^ Three papers explored self-described coping strategies.^36,37,41^

**Table 3.** Assessments and definitions of different concepts of ‘negative emotions’ and coping mechanisms related to climate change and mental health

| **NEGATIVE EMOTIONS** |  |
| --- | --- |
| **Concerns** |  |
| Agho et al. (2010)<sup>34</sup> | “If global warming were to worsen, how concerned would you be that you or your family would be directly affected by It?” |
| Strife et al. (2012)<sup>38</sup> | “Environmental concern referred to the kinds of environmental problems (if any) the children deemed important” |
| El Zoghbi & El Ansari (2014)<sup>40</sup> | Concerns over inequalities in impacts of climate change and adaptation mechanisms, evaluated in the qualitative interview. |
| Li & Monroe (2019)<sup>42</sup> | Concern about climate change measured using one question. However, it is unclear how the question was framed. |
| **Worry** |  |
| Henker, Whalen & O’Neil (1995)<sup>31</sup> | Qualitative evaluation via asking students “Everybody worries some of the time. What are some things that you worry about-like things that you think hard about, or think about lot” |
| Ojala (2005)<sup>32</sup> | Level of worry about different environmental risks including climate change. |
| Ojala (2012a)<sup>36</sup> | “How much worry you felt about climate change?” |
| Ojala (2012b)<sup>37</sup> | Five questions on how much the children worried about negative consequences caused by climate change for themselves, their close ones, future generations, people living in economically deprived countries, and animals/nature |
| **Other negative emotions associated with climate change** |  |
| Searle & Gow (2010)<sup>35</sup> | “Thinking about climate change now makes me feel – concerned, tense, worried, anxious, depressed, hopeless, powerless, sad, helpless, stressed, angry, scared”. |
| Strife et al. (2012)<sup>38</sup> | “How does this environmental concern make you feel?” |
| MacDonald et al. (2015)<sup>41</sup> | Worry, boredom, loneliness, isolation, restlessness, fear, sadness, anger, frustration, stress, anxiety, alcohol and substance abuse related to the changing local environment |
| **MENTAL HEALTH AND WELLBEING OUTCOMES** |  |
| **Mental ill-health** |  |
| Wolf & Sala (2008)<sup>33</sup> | Clinical diagnosis of Major Depressive Disorder with psychotic features |
| Searle & Gow (2010)<sup>35</sup> | Depression, anxiety and stress measured by DASS-21 |
| Searle & Gow (2010)<sup>35</sup> | General state of apprehension, fear, worry and concern about unfavourable changes in a more remote personal future measured by the Future Anxiety Scale (FAS) |
| Ojala (2012b)<sup>37</sup> | Anxious and depressive feelings felt during the last week using the Child Depression Scale |
| MacDonald et al. (2015)<sup>41</sup> | Whether the participant struggled with any mental health-related issues or challenges at any time in their lives, whether they accessed mental health services |
| **Well-being** |  |
| Ojala (2005)<sup>32</sup> | Subjective well-being on coping, energy level, physical and emotional condition. |
| Ojala (2012b)<sup>37</sup> | Subjective life satisfaction using a seven-item scale measuring factors such as “I have a good life”, and “I wish I had a different kind of life” |
| **BEHAVIOURAL OUTCOMES** |  |
| Agho et al. (2010)<sup>34</sup> | “How much have you changed the way you live your life because the possibility of worsening global warming?” |
| Ojala (2012b)<sup>37</sup> | Pro-environmental behaviour measured using twelve items capturing both behaviour in everyday life such as recycling, ride to school and influencing others |
| **COPING STRATEGIES** |  |
| **Efficacy** |  |
| Ojala (2012b)<sup>37</sup> | Measured using 4-item instrument on individual self-efficacy and collective efficacy.<br>• “I think that I myself can contribute to the improvement of the climate change situation”<br>• “I know there are a number of things that myself can do in order to improve the climate change problem”<br>• “I believe that together we can do something about the climate threat”<br>• “I am confident in that we together can solve the climate change problem”. |
| Li & Monroe (2019)<sup>42</sup> | Efficacy was defined as the belief that society and lay people have the ability and skills to undertake actions. It was measured using a 4-item scale:<br>• “I believe that people will be able to fix climate change<br>• “I believe that research and technical solutions will help fix climate change”<br>• “Forest landowners can make a difference in the climate by practicing good forest management strategies”<br>• “Because people can change their behaviours, we can influence climate change in a positive direction” |
| **Hope** |  |
| Ojala (2012a)<sup>36</sup> | “How much hope you felt about climate change” |
| Ojala (2012b)<sup>37</sup> | Optimism concerning climate change measured by three items:<br>• “I feel hopeful that we will fix the climate change problem in the future”<br>• “I think we will solve the climate problem in the future”<br>• “I believe the future looks bright when it comes to climate change”. |
| Strife et al. (2012)<sup>38</sup> | Evaluate children's perceptions of their future with questions on what they thought the earth would look like in 100 years and what their hopes and worries were about the future. |
| Li & Monroe (2019)<sup>42</sup> | Agency and pathways thinking about hope of climate change<br>• "I am hopeful about resolving climate change because more people are taking it seriously"<br>• "I know that there are a number of things that I can do to contribute to climate change solutions"<br>• "I am hopeful about climate change because I can think of many ways to resolve this" |
| **Coping** |  |
| Ojala (2012a)<sup>36</sup> | "When you feel the most worried, do you do any-thing to not worry so much? If yes, describe what you do?" |
| Ojala (2012b)<sup>37</sup> | Themes were defined based on response included problem-focused, emotion-focused (de-emphasizing) and meaning-focused coping |
| MacDonald et al. (2015)<sup>41</sup> | Coping mechanisms including access to land as well as family and community support |

Only two studies evaluated psychological symptoms, namely depression and stress, using validated psychometric scales.^35,37^ One study conducted a survey with Swedish young people (n=293), and found that negative affect measured by the Child Depression Scale was positively correlated with problem-solving coping, but negatively correlated with meaning-focused coping. Meaning-focused coping was also positively related to life satisfaction and general positive affect. The second study is the only one that directly examined the link between psychological symptoms and worry about the threat of climate change. An Australian survey (n=275) measured symptoms indicative of clinically significant emotional states of depression, anxiety, and stress using the DASS-21, and found higher scores for respondents who reported higher climate change distress.^35^ The study included a mix of age groups, with young adults age 18-25 years making up a majority (57%) of the sample, and demonstrating the highest climate change distress and anxiety. One qualitative study reported whether participants had ever struggled with any mental health-related issues or challenges at any time in their lives, but did not relate this to climate anxiety.^41^

### Impact of specific climactic events on young people’s mental health

During the literature search it was clear that most of the research on mental health and climate change conducted to date related to specific climatic events. An exploratory article count was performed and articles that examined the effect of natural disasters on youth mental health were mapped across time from first publication to present (Figure 2).

**Figure 2.**
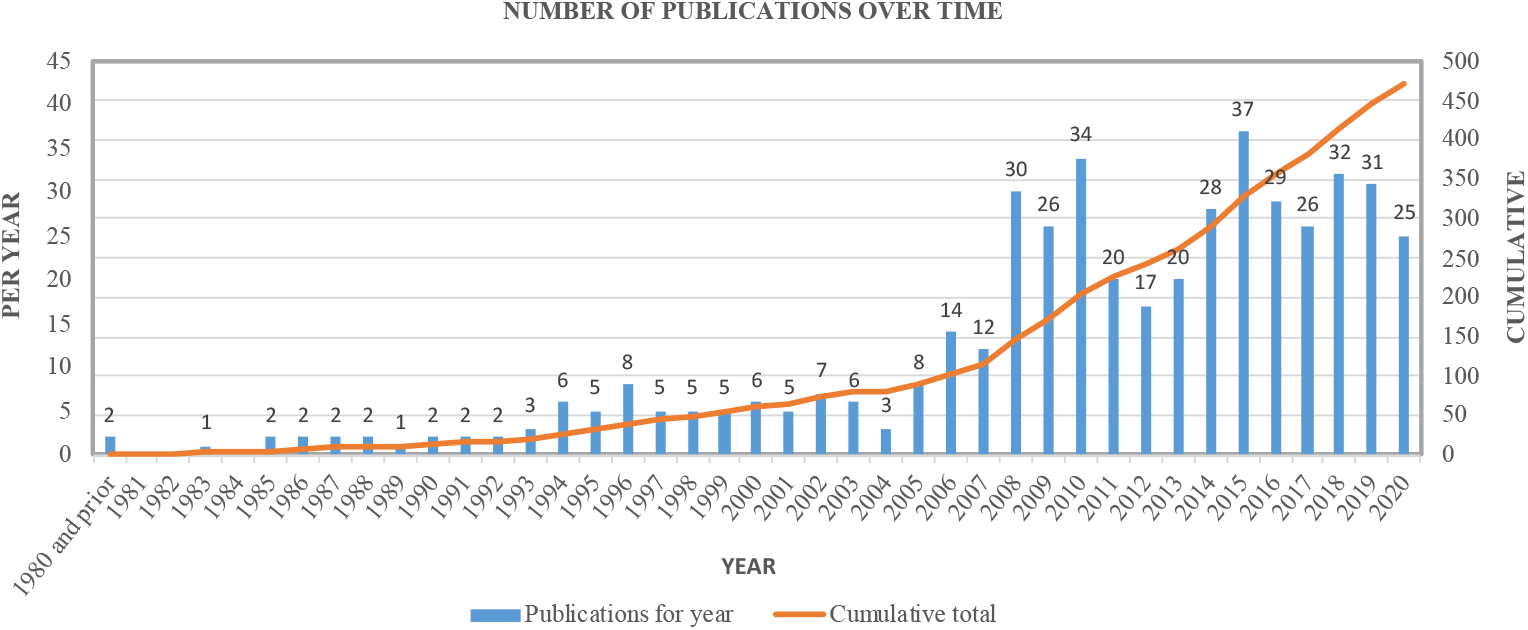
Number of journal articles that discuss the mental health of young people in relation to a natural disaster published annually over time

## DISCUSSION

The present review provides an overview of the current evidence for how young people experience negative emotions towards climate change. To the best of our knowledge, the present review represents the first systematic review of the evidence related to climate anxiety in young people. Overall, evidence is scarce, with most research conducted in the last 10 years. Although the causal relationship between worry about climate change and a diagnosis of mental illness was not directly examined in any included papers, available evidence suggests that young people are worried and experienced a range of negative emotions about the threat of current and future effects of climate change.

Although only one study directly examines the connection between climate distress and clinically significant symptoms of depression, anxiety and stress,^35^ the findings suggest a possible link between experiences of climate anxiety and mental health in young people, and give support to the idea that young people may be at heightened risk of mental health impacts compared to older people.

Several reasons have been proposed to explain young people’s vulnerability to the negative impacts of climate anxiety, including neurobiological changes occurring in the brain.^28^ Indeed, young people are already at heightened risk of emerging mental ill-health compared to other age groups, with prevalence of mental illness among young people being the highest of any age group.^43^ Across several studies included the present review, young people reported concern about climate change not only in the present but also into the future. This is consistent with the view of climate anxiety as owing to an “existential” threat to life itself.^9,10^ Research on other existential global threats, such as the fear of nuclear war, have found that this type of concern is associated with an increased risk for common mental disorders in adolescents.^44^ This further supports the argument that young people are likely to be at higher risk for the detrimental effects of climate-related worry. While lack of data currently makes it difficult to draw firm conclusions about the mechanisms or long-term mental health impacts of climate-related worry in young people, there is a clear argument for youth being a critical period both for emergence and prevention of climate-related mental health problems.

Positively, results of the current review also revealed evidence for factors that may help to promote well-being among young people in the face of climate change. This was an unexpected finding, as our research question and search strategy aimed at understanding the evidence related to mental health and negative emotions associated with climate anxiety, rather than how young people cope with existential threat. Results suggest a number factors that can support wellbeing or coping related to climate change, including meaningfulness, hope, and optimism regarding climate change, an awareness of possible solutions, a high degree in trust in environmental organisations, and motivation to engage in positive environmental behaviours.^27,32,37,38,42^ Findings of one research group showed that different types of coping strategies can be more or less effective for reducing negative affect and promoting well-being.^32,36^ Specifically, meaning-focused coping (such as the belief that “more and more people have started to take climate change seriously”) was associated with improved wellbeing and reduced negative affect, while problem-focused coping such as “thinking about what I myself can do”) was associated with higher negative affect.^37,39^ Findings from this group also showed that worry about the environment could co-occur with both positive and negative wellbeing, suggesting that coping may act to support wellbeing without impacting level of worry. This is consistent with the idea that negative feelings associated with climate change are not necessarily pathological in and of themselves.^10,45^ In fact, anxiety can act as a source of efficacy beliefs and motivation for change.^46^ However further research is needed to understand underlying mechanisms in larger and diverse samples.

Although not the focus of any included studies, another way that young people may be coping with climate change is through activism. Recent youth-led movements have been attended by millions of young people across 150 countries. Children and adolescents have also sued their government in Australia,^47^ Canada,^48^ the United States,^49^ and several other countries^50^ for their inaction in response to the climate crisis. While there is a body of literature suggesting that engaging in political activism can have positive psychological and social outcomes for young people,^51^ there is also evidence to suggest that major protests may increase the prevalence of depression, even without direct involvement.^52^ In this review the authors posit that this is due to disruptions in interpersonal relationships related to ideological disputes following the protests. While coping is widely-accepted as a key concept for understanding the impact of negative emotions such as worry on mental health and well-being, there are at least 100 taxonomies of coping proposed in the literature. Future research on this and other types of coping could help build understandings of who might benefit from different strategies to support wellbeing, and why.

Figure 3 provides a conceptual framework of how negative emotions associated with the existential threat of climate change may interact with and impact young people’s mental health. Negative emotions such as anxiety, concern, worry or despair, while not pathological by themselves, may increase the risk of mental ill-health if they become prolonged and overwhelming. While the evidence for the long-term mental health risks associated with climate anxiety is scarce, there is a strong body of research to show that other types of long-term stress increases the risk of mental health problems such as depression, anxiety, and substance use problems. In the context of climate change, the risk of emergence of mental ill-health may be increased by individual or collective risk factors, including the direct effects of climate change such as displacement and exposure to traumatic climatic events and other predisposing risk factors such as pre-existing physical or mental ill-health. This risk may be buffered by healthy coping strategies, such as meaning-focused strategies, and by protective factors like having a strong connection to culture.

**Figure 3.**
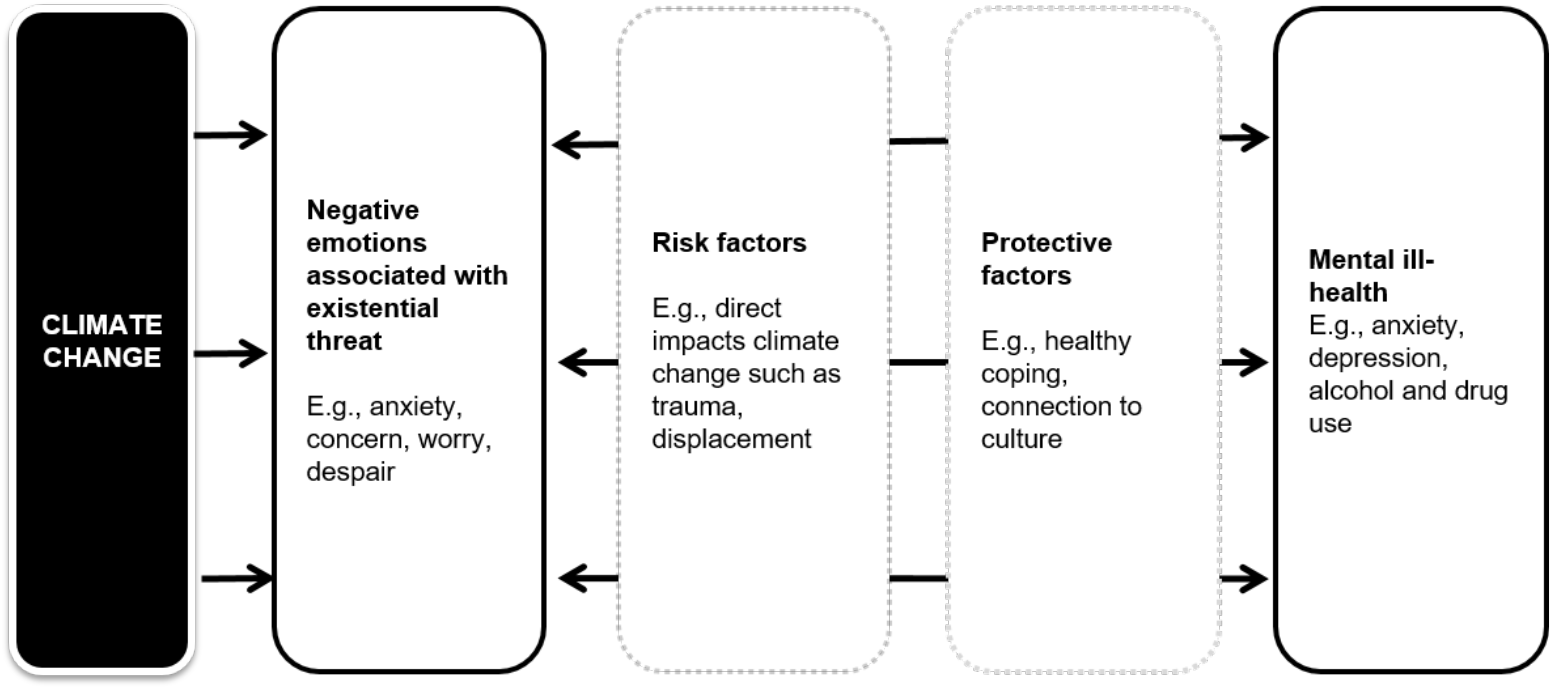
Conceptual framework showing the interaction between negative emotions associated with the existential threat of climate change and mental ill-health. Risk and protective factors may act to mediate risk of negative emotions contributing to the development of mental ill-health

The small number of studies included in this review were heterogeneous in design. Some used qualitative methods (mainly interviews and focus groups), others quantitative (surveys). Most of the studies did not aim to evaluate the prevalence or severity of concerns and negative emotions related to climate change, and no study evaluated how these negative emotions may increase risk of psychiatric morbidity. Two studies considered existing mental health issues of participants, but did not explore differences in the experience of climate-related distress between those with existing mental health problems compared to those without. In addition, no studies looked at climate-related worry over time to explore population-level risk and protective factors or changes at the individual level. Measures used across studies to investigate climate anxiety and related experiences were inconsistent, apart from those conducted by the same author group. Variation in terms and measures is consistent with wider literature in the area of climate change and mental health, which includes a wide range of terms to describe these concepts (see Table 1). Although, the term “climate anxiety” has been widely used, there is currently a lack of consensus on what aspects of negative emotions it should cover, and how its severity should be measured. This makes it difficult to compare the overall levels of concerns and negative emotions across studies in this review and across the field.

The majority of included studies were conducted in industrialised countries with largely White populations, with 10 out of 12 studies conducted in three countries (Sweden, Australia, and USA). The limited diversity in study settings and populations is consistent with findings of other reviews on the impact of climate change on health^4^ and further adds to the clear need for more research in this area overall. Only one qualitative study included in the present review was conducted with an Indigenous community.^41^ Findings of that study suggest that there may be unique factors that help to support mental health and wellbeing of First Nations young people in the face of climate change, such as the protective power of being on the land and connecting to Indigenous culture.

The relatively small number of studies that met inclusion criteria for this review contrasts with the large amount of research found on the direct impact of natural disasters on the mental health of young people, which included nearly 500 peer-reviewed articles to-date. Visual analysis of trends over time (Figure 2) shows that research in this area was extremely scarce until the mid-1980s. Following 2005, Hurricane Katrina prompted a spike in publications, and publications per year have been relatively stable since. The increasing number of publications examining the effect of natural disaster on mental health reflects the increasing frequency of natural disasters worldwide^53^ and the growing understanding of their direct effect on human health and wellbeing.^50^ It is also representative of the increased understanding of the impact of distressing experiences on the development of children and young people.^54^ Main findings of the present review highlight the need for more research that considers the experiences of young people who have not been directly impacted by a specific climatic event as well as those who have. As the frequency of climate disasters increases, the likelihood that more people will be directly impacted by such events also increases. Understanding how climate-related worry may interact with stress of experiencing climatic events is a key area for further investigation.

The scant evidence around climate anxiety in young people is seemingly at odds with available evidence from grey literature that suggests many young people are worried about climate change. A recent survey of more than 2000 young people in the UK found that almost three quarters (73%) said they are worried, and one in five (22%) said they were very worried about the state of the planet.^55^ Australia’s largest online youth survey of 15 to 19 year-olds (n=25,126^25^ and n=25,800^26^) has similarly placed mental health and the environment amongst the top four most important issues to young people for the last two years. Large-scale, rigorous surveys could add to these findings and paint a picture of the prevalence of climate concern among the population.

### Implications for research and clinical practice

Findings clearly show that experiences of climate anxiety in young people is understudied, with only 12 articles included through a systematic literature search and screen of over 3000 articles. This adds support to Wu and colleagues’ call to action to researchers, mental health professionals and policy-makers to better understand the effects of climate anxiety on youth mental health in order to improve the mental health and wellbeing of our future leaders.^28^ Further research is needed to characterise climate concern in young people, to clarify if there are qualitative and/or quantitative differences between adults and young people in experiences of climate-related worry and coping. Longitudinal research is needed to assess climate change-related negative emotions and mental health outcomes over time. Such research would offer invaluable data on potential factors contributing to vulnerability and/or resilience towards the development and trajectory of psychological impact of climate change.

To support comparison and harmonisation across studies, structured measures of climate anxiety must be developed and validated. It is currently unclear if climate anxiety is qualitatively or quantitatively different across the lifespan or across cultural groups. This may be particularly relevant for First Nations Peoples, who have strong cultural connection with nature and the land, as well as other historically marginalised groups who may have experience with existential threat to community and culture from human actions such as colonisation. More broadly, more research is needed with young people from a range of diverse backgrounds, particularly while the language and thinking around climate-related worry and anxiety is still emerging. To avoid creating definitions that centre the experience of White, wealthy communities, researchers must engage in genuine collaboration with young people with diverse experiences and identities to design research, and develop terminology and measures that consider the complexity of climate anxiety as a phenomenon. There is an urgent need to develop evidence-based conceptual frameworks defining climate anxiety and the relationship to mental health and wellbeing, along with robust measures that capture different dimensions and severities of the issue for different populations.

Results of this scoping review also have implications for youth mental health care. Prevention and intervention initiatives could aim to target coping mechanisms, in particular those shown to support wellbeing and reduce negative affect (i.e., meaning-based coping). Education programs and materials may be one strategy for helping to build the capacity and confidence of young people to reduce their own footprint and advocate for other individuals and organisations to take action. Given the large numbers of young people reporting worry about climate change, we suggest that mental health professionals ask about climate-related worry as part of their routine clinical assessment to inform approaches to care.

In addition, the mental health system itself needs to be resilient and responsive to the challenges brought by climate change, through the development of risk assessment and adaptation plans that respond to climate change through the building of climate-resilient health infrastructure and workforce.^56^ Mental health services can also play a direct role in climate change mitigation by reducing their greenhouse emissions through the adoption of environmentally sustainable practices and infrastructure.^56^ Beyond research and clinical practice, young people are concerned over lack of action by the governments and policy-makers.^47-50^ Policy inaction on climate change will threaten the mental well-being and lives of individuals and communities.^57^ A recent survey of 10,000 young people in ten countries showed a profound impact of climate change on young people’s psychological and emotional reactions, which was associated with their perceptions of governments’ responses to climate change.^58^ Thus, arguably the most important role mental health leaders and service providers can play is to show leadership in advocating for climate change action consistent with the concerns of young people. The cost of inaction is also disproportionately greater for those who are most affected by climate change; therefore, climate change mitigation is essential for climate and intergenerational justice as well as the prevention of mental health consequences of climate change.^59,60^

### Strengths and limitations

The main strength of this review is that it was created by performing comprehensive systematic literature searches of several major databases using carefully determined and refined terms and selection criteria. A subset of papers was double-screened and all included papers were discussed by the author group, which includes researchers with prior experience screening and synthesising peer-reviewed literature. However, only papers published in English, French, or German were included and searches did not include grey literature, which means that some relevant evidence may have been missed and the cultural and geographic diversity of included papers may have been limited. In addition, the quality of included papers was not rigorously assessed, hence conclusions made should be interpreted with some caution.

## CONCLUSION

Building the evidence-base around the mental health and wellbeing impacts of the threat of climate change in young people is an urgent priority. Understanding the evidence for the impact of the threat of climate change on the mental health of young people is essential for designing interventions and approaches that aim to improve outcomes for young people experiencing distress. Future research should seek to measure the relationship between climate anxiety and mental health outcomes over time in people with various cultural and socioeconomic identity, and to further build knowledge of protective factors and coping mechanisms. In the meantime, governments and mental health professionals can support youth mental health by acknowledging the real threat of climate change to young people’s wellbeing and by engaging in action to mitigate the effects of climate change.

## Data Availability

There is no data associated with this study.

## Supplementary Material

### Database Searches

#### EMBASE (Ovid)

| # | Query |
| --- | --- |
| 1 | climate change/ or greenhouse effect/ or sea level rise/ or hurricane/ or tornado/ or drought/ or flooding/ or wildfire/ or extreme hot weather/ or extreme weather/ or sea water/ or ice cover/ or species extinction/ or vector borne disease/ |
| 2 | (climate change or climate crisis or global warming).ti,ab. |
| 3 | (climat* adj3 (heating or variabilit* or emergenc*)).ti,ab. |
| 4 | (greenhouse adj3 (effect* or gas or gasses)).ti,ab. |
| 5 | ((extreme or disaster*) adj3 (weather or climati*) adj3 (event* or phenomen*)).ti,ab. |
| 6 | (heat wave* or surface air temperature*).ti,ab. |
| 7 | ((rise or rising) adj3 temperature*).ti,ab. |
| 8 | (floods or flooding or hurricane* or tornado* or drought or wildfire* or bush fire* or deglaciation or deforestation or desertification or vector borne disease*).ti,ab. |
| 9 | (solastalgia or ecoanxiety or eco-anxiety or ecomigrat* or eco-migrat*).ti,ab. |
| 10 | or/1-9 |
| 11 | adolescent/ or young adult/ or student/ or child/ |
| 12 | (young people or youth* or young or adolescen* or student* or pubescen* or teenage* or undergraduate* or young men or young women or young adult* or juvenile* or child*).ti,ab. |
| 13 | or/11-12 |
| 14 | mental health/ or exp mental disease/ or mental stress/ or distress syndrome/ or exp mood disorder/ or exp drug dependence/ or exp eating disorder/ or suicide/ or exp automutilation/ or psychological resilience/ or exp coping behavior/ |
| 15 | (mental health or mental illness or mental disorder* or mental ill-health or wellbeing or well-being or stress or distress or psychological or psychiatric or depressi* or mood disorder* or dysthymi* or major depressive disorder* or affective disorder* or anxiety or panic or phobia or acute stress disorder* or post-traumatic stress or posttraumatic stress or obsessive compulsive disorder* or "alcohol and other drugs" or "substance use" or "alcohol use" or "drug use" or alcohol abuse or drug abuse or substance misuse or drug misuse or alcohol misuse or substance dependenc* or drug dependenc* or alcohol dependenc* or alcohol addiction or substance addiction or drug addiction or bipolar disorder* or cyclothymi* or eating disorder* or anorexia or bulimia or purging or binge eating or "eating disorder not otherwise specified" or "feeding and eating disorder*" or psychosis or psychotic or delusion or hallucination or paranoi* or schizopreni* or suicid* or self-harm or self-injur* or self-mutilat* or self-destructive or automutilation or self-poisoning or self-inflict* or resilience or adaptation).ti,ab. |
| 16 | or/14-15 |
| 17 | 10 and 13 and 16 |
| 18 | limit 17 to (article or article in press) |

#### Medline (Ovid)

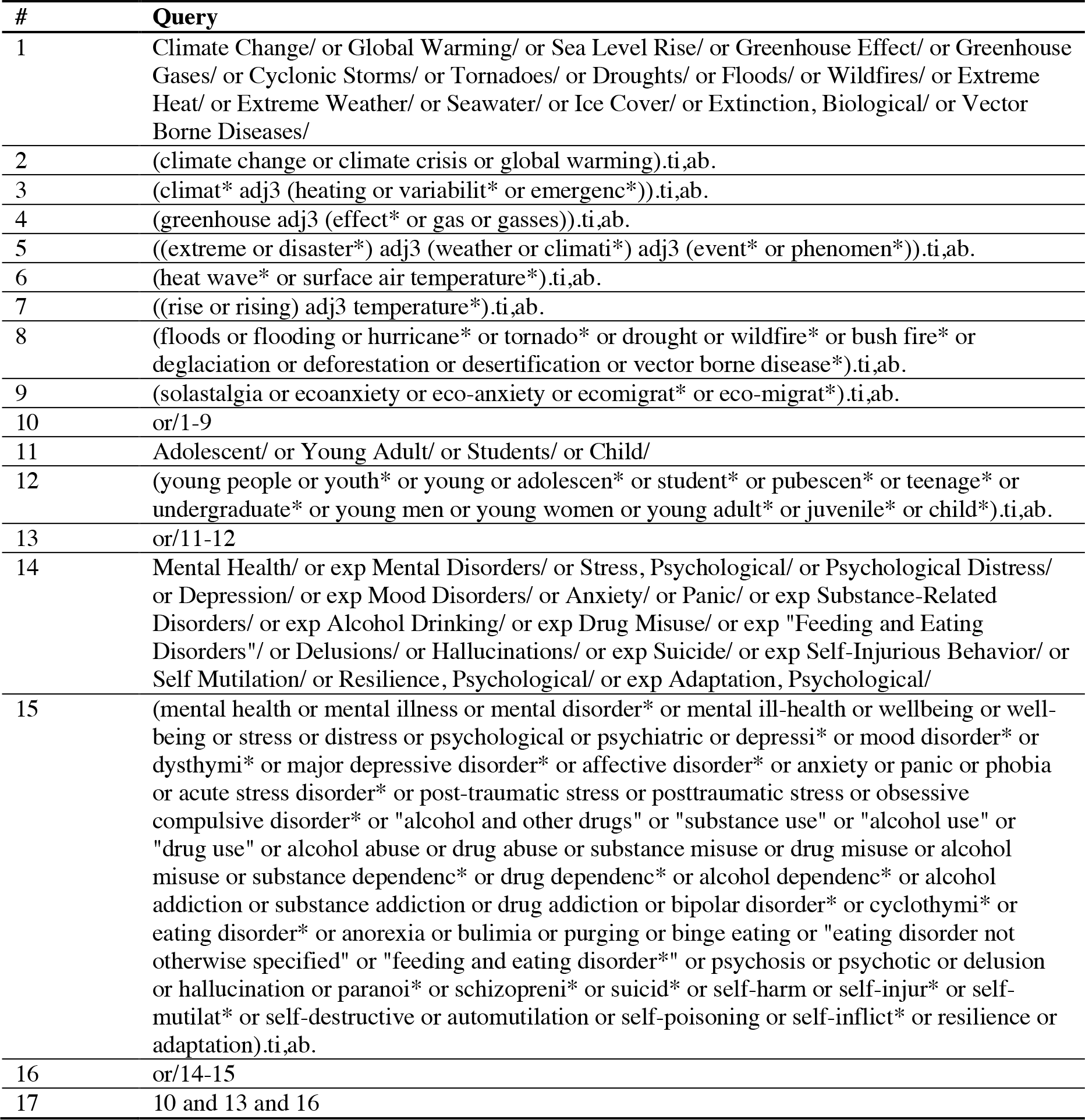

#### PsycINFO (Ovid)

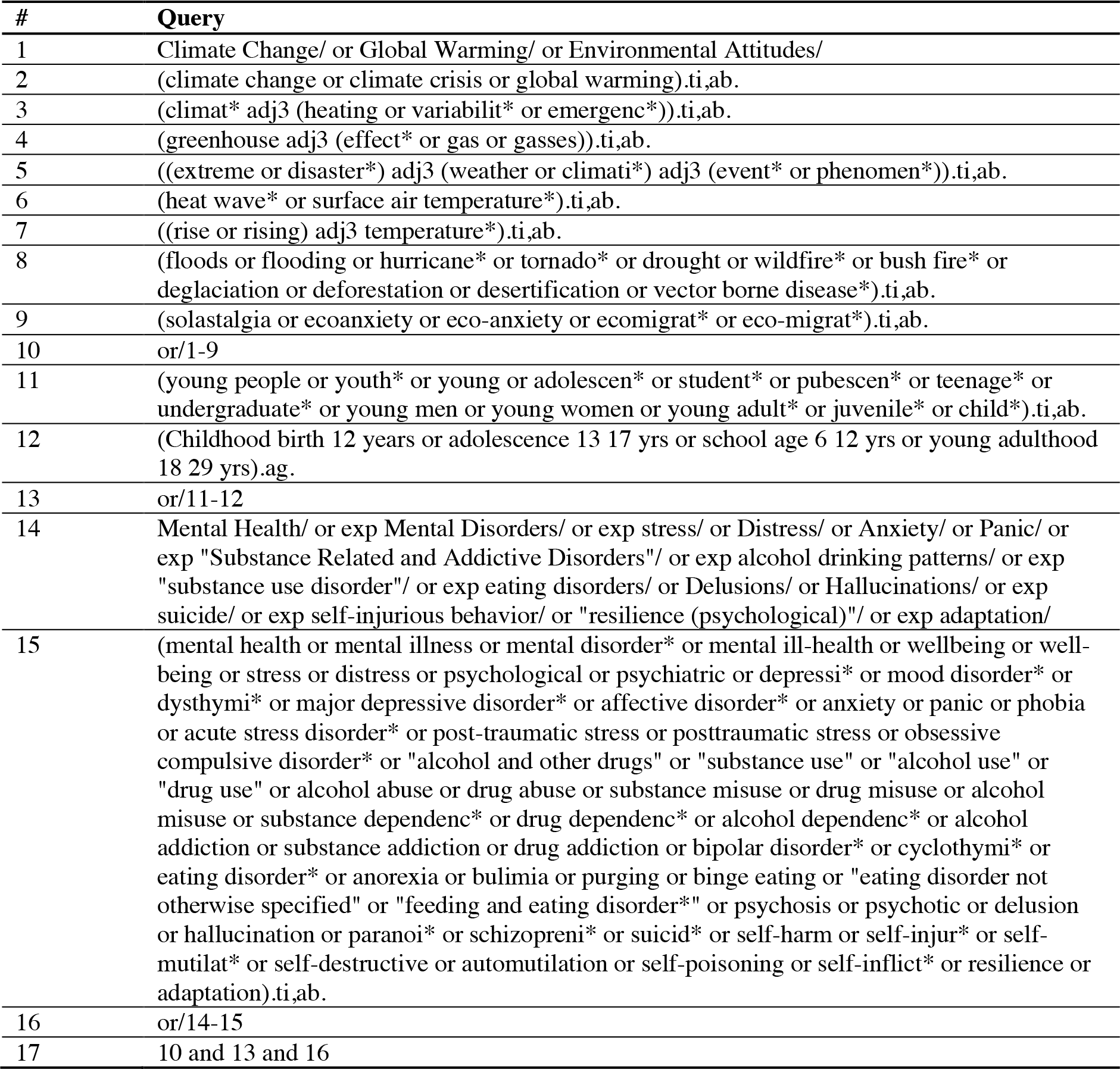

| # | Query |
| --- | --- |
| 1 | Climate Change/ or Global Warming/ or Environmental Attitudes/ |
| 2 | (climate change or climate crisis or global warming).ti,ab. |
| 3 | (climat* adj3 (heating or variabilit* or emergenc*)).ti,ab. |
| 4 | (greenhouse adj3 (effect* or gas or gasses)).ti,ab. |
| 5 | ((extreme or disaster*) adj3 (weather or climati*) adj3 (event* or phenomen*)).ti,ab. |
| 6 | (heat wave* or surface air temperature*).ti,ab. |
| 7 | ((rise or rising) adj3 temperature*).ti,ab. |
| 8 | (floods or flooding or hurricane* or tornado* or drought or wildfire* or bush fire* or deglaciation or deforestation or desertification or vector borne disease*).ti,ab. |
| 9 | (solastalgia or ecoanxiety or eco-anxiety or ecomigrat* or eco-migrat*).ti,ab. |
| 10 | or/1-9 |
| 11 | (young people or youth* or young or adolescen* or student* or pubescen* or teenage* or undergraduate* or young men or young women or young adult* or juvenile* or child*).ti,ab. |
| 12 | (Childhood birth 12 years or adolescence 13 17 yrs or school age 6 12 yrs or young adulthood 18 29 yrs).ag. |
| 13 | or/11-12 |
| 14 | Mental Health/ or exp Mental Disorders/ or exp stress/ or Distress/ or Anxiety/ or Panic/ or exp "Substance Related and Addictive Disorders"/ or exp alcohol drinking patterns/ or exp "substance use disorder"/ or exp eating disorders/ or Delusions/ or Hallucinations/ or exp suicide/ or exp self-injurious behavior/ or "resilience (psychological)"/ or exp adaptation/ |
| 15 | (mental health or mental illness or mental disorder* or mental ill-health or wellbeing or well-being or stress or distress or psychological or psychiatric or depressi* or mood disorder* or dysthymi* or major depressive disorder* or affective disorder* or anxiety or panic or phobia or acute stress disorder* or post-traumatic stress or posttraumatic stress or obsessive compulsive disorder* or "alcohol and other drugs" or "substance use" or "alcohol use" or "drug use" or alcohol abuse or drug abuse or substance misuse or drug misuse or alcohol misuse or substance dependenc* or drug dependenc* or alcohol dependenc* or alcohol addiction or substance addiction or drug addiction or bipolar disorder* or cyclothymi* or eating disorder* or anorexia or bulimia or purging or binge eating or "eating disorder not otherwise specified" or "feeding and eating disorder*" or psychosis or psychotic or delusion or hallucination or paranoi* or schizopreni* or suicid* or self-harm or self-injur* or self-mutilat* or self-destructive or automutilation or self-poisoning or self-inflict* or resilience or adaptation).ti,ab. |
| 16 | or/14-15 |
| 17 | 10 and 13 and 16 |

